# THE RELATIONSHIP BETWEEN CLINICAL PROFILE, GLYCEMIC PARAMETERS AND HYPOGLYCEMIA IN TYPE 1 DIABETES PEDIATRIC PATIENTS

**DOI:** 10.64898/2025.12.19.25342708

**Authors:** Andreea Morar-Stan, Luminiţa Dobrotă, Anişoara Răduţu, Carmen Daniela Domnariu

## Abstract

**Background/Objectives:** Our objective was to understand the role of clinical and continuous glucose monitoring (CGM) parameters in predicting the risk of hypoglycemia in type 1 diabetes pediatric patients.

**Methods:** Pediatric patients with type 1 diabetes (n=71) at the Oradea County Clinical Emergency Hospital, Romania, underwentCGM during their initial visit and were followed for at least 6 months, with in-clinic visits every 3 months. Age, body mass index, time in range, mean daily glucose(MDG) concentration, coefficient of variation(%CV)were considered as potential predictors of the risk of hypoglycemia (presented as the percentage of time spent below two glycemic thresholds: 3.9 and 3.0 mmol/L, corresponding to mild, respectively clinically significant hypoglycemia).

**Results:** Looking at a total of 142 glycemic profiles, MDG was significantly lower in those with hypoglycemia compared to those without, while %CV was significantly higher(p<0.0001). Regression tree models identified %CV as the dominant variable for both thresholds, while classification tree models identified %CV for clinically significant hypoglycemia and MDG for mild hypoglycemia. It was observed that in profiles with a %CV below 36.15% and MDG above 7.16 mmol/L, the mean percentage of time spent below the 3.9 mmol/L threshold was 4.8%—a value close to that recommended in American Diabetes Association guidelines. Patients younger than 7 years presented the highest frequency of both mild and clinically significant hypoglycemic episodes.

**Conclusions:** Our study supports %CV and MDG concentration as key factors in predicting hypoglycemia risk. Minimizing the risk of hypoglycemia in pediatric patients requires a %CV below 36%.

## 1. Introduction

Type 1 diabetes mellitus (T1DM), known as the most prevalent form of diabetes in the pediatric population, is a chronic autoimmune condition that leads to the progressive destruction of pancreatic β-cells and, consequently, to an absolute insulin deficiency [1,2].The continuous global rise in T1DM incidence represents a major clinical concern, as it has become one of the leading endocrine disorders of childhood [3,4]. This epidemiological reality requires the adoption of strict glycemic management based on intensive exogenous insulin replacement therapy, aiming to increase the time spent in euglycemia and to minimize episodes of hypoglycemia and hyperglycemia. However, achieving glycemic targets is often suboptimal due to a convergence of age-specific and developmental factors, all contributing to a reduced quality of life for both pediatric patients and their caregivers [5,6]. Given the inherent instability of glycemic control, recurrent hypoglycemic episodes remain a persistent risk, their clinical relevance in pediatrics being largely attributable to their effects on cognitive functioning—an area extensively documented in the literature [7–12]. Studies indicate that cognitive function returns to baseline within 40–90 minutes after euglycemia is restored [13]. Nevertheless, concerns persist regarding the long-term effects of hypoglycemia on cognition, particularly among patients with early-onset disease.

In response to these challenges, technological innovations, including continuous subcutaneous insulin infusion (CSII) pumps and continuous glucose monitoring (CGM) systems, have emerged. Their clinical integration has been associated with reduced hypoglycemia and substantially improved glycemic control [14–17].

The therapeutic approach to individuals with type 1 diabetes, from a glucocentric perspective, requires close monitoring of several key points: ambient hyperglycemia, time spent within the target range [18,19], short-term glycemic variability [20–22], and hypoglycemia thresholds [23,24]. In this context, the use of Ambulatory Glucose Profile (AGP) reports in pediatric clinical practice has proven to be a feasible option, allowing continuous assessment of glycemic status and, implicitly, the identification of hypoglycemic episodes that may otherwise remain undetected during routine clinical care [25]. According to the International Consensus on the Use of CGM, among the parameters quantified using AGP, the coefficient of variation (%CV) is considered a key indicator distinguishing stable from unstable diabetes, with superior sensitivity for detecting hypoglycemic excursions. Beyond a %CV of 36%, the frequency of hypoglycemia is significantly increased, particularly in insulin-treated patients [26–28]. Alongside %CV, mean daily glucose (MDG) appears to be another major explanatory factor contributing to hypoglycemia risk. However, only a few studies have investigated the appropriateness of these thresholds in pediatric patients, with this being the first one conducted in Romania.

In this observational and retrospective study, using data from AGP in children and adolescents with type 1 diabetes, we aimed to identify the role of %CV and MDG in determining the time spent below the thresholds of 3.9 mmol/L and 3.0 mmol/L, values that represent hypoglycemia [29].

## 2. Materials and Methods

We conducted a retrospective observational study using data from patients with T1DM admitted to the Department of Diabetology and Nutrition Diseases, Pediatric I of Oradea County Clinical Emergency Hospital. The main objective of this study was to analyse the role of clinical and CGM parameters in determining the time spent below the thresholds of 3.9 mmol/L and 3.0 mmol/L, values that represent hypoglycemia.

The present study was approved by the Ethics Committee of the Oradea County Emergency Clinical Hospital (No. 19384/ 26 June 2025/) and the “Lucian Blaga” University of Sibiu (No.57/12 November2025). The present study is in accordance with the ethical principles set out in the Declaration of Helsinki.

Inclusion criteria were: type 1 diabetes diagnosis according to the American Diabetes Association classification, age between 2-18 years, users of real-time CGM (rtCGM) systems, a well-defined treatment regimen in the last 3 months, parental consent, and willingness and ability to adhere to the study protocol, along with an information system that met the requirements for uploading study data [30]. Exclusion criteria for analysis were: other forms of diabetes, absence of data for any of the studied variables, unexpected interruption of glucose monitoring, corticosteroid therapy in the last 3 months, and pregnancy.

Patients were asked to wear a rtCGM device (Guardian™4 sensor) during their initial visit and were followed for at least 6 months, with in-clinic visits every 3 months. RtCGM data were uploaded at every visit. For each patient, two AGPs were obtained over a six-month interval. Consequently, the final analysis sample included 142 profiles. For data from subjects to be included in the analyses, at least 90 days of consecutive CGM wear and sensor wear time ≥70% was required based on previous guidelines [31–33].

The %CV, MDG, time in range (TIR) and the time spent below two glucose thresholds (3.9 mmol/L and 3.0 mmol/L) were the main variables involved in our study.

Wilcoxon rank sum test with continuity correction was conducted using the Wilcox.test function in order to compare means between two groups.

Univariate linear mixed models were used to explore the existence of an association between explanatory variables: age, body mass index (BMI), MDG, %CV, TIR and insulin dose, and the response variables: percentage of time spent below glucose thresholds 3.0 and 3.9 mmol/L. These were chosen as two glycemic profiles were available per participant, and ran using the lmer command (e.g. lmer(arcsin(sqrt(percentage of time spent below threshold/max(percentage of time spent below threshold)) ~ age + (1|participant), data=dataset)) from the lme4 package. The function Anova from the car package was used to conduct a Type II Wald chi-square test to assess the significance of each explanatory variable.

Recursive Partitioning and Regression Trees were fitted using the function rpart from package rpart. All variables that were significant in the univariate linear mixed models analysis were included as explanatory variables, while the response variable were: percentage of time spent below glucose thresholds 3.0 (clinically significant hypoglycemia), and 3.9 mmol/L (mild hypoglycemia) (regression), any time versus no time spent below each glucose thresholds (classification). For each tree, a complexity parameter table was plotted using the plot cp function and the tree depth corresponding to the first value on the left of the plot that lies underneath the horizontal line was chosen.

Beta regression was used to model the relationship between the percentage of time spent below the glucose thresholds and MDG, respectively %CV. This type of regression is specifically designed to analyse continuous variables that are bounded between 0 and 1, and avoids unrealistic predictions outside this range. Percentages were divided by 100 and 0.001 was added to all of these as the Beta distribution does not include 0. The constant 0.001 was then subtracted from the predictions before multiplying them by 100.These models were fitted using the betareg package. For comparison and following a previous study [34], a linear model was also fitted on the log-transformed data after the above transformations were also applied as the natural logarithm is undefined for 0.

To examine whether there is a relationship between age category (<7, 7-13, >13 years) and the risk of mild and clinically significant hypoglycemic events we created two-way contingency tables and conducted the Chi-Square test of independence, testing for evidence against the null hypothesis of no relationship (independence).

## 3. Results

Our study had a total of 71 participants, with two glycemic profile measurements each (142 glycemic profiles), of whom 32 were receiving continuous subcutaneous insulin infusion (CSII). Ages ranged from 3 to 18, median 13 (interquartile range (IQR) 9-15), 36 (51%) males, BMI from 14.10 to 31.10, median 20.00 (IQR 18.30-22.05), 6 (8%) patients had had diabetes for less than a year, 31 (44%) between 1-5 years, and 34 (48%) for more than 5 years. HbA1C had a mean 7.9, standard deviation (SD) 1.9, and range 5.3-15.1 (n=71), MDG 8.1, 1.2, 5.9-13.4 (n=142), %CV 39.1%, 6.1%, 23.8–61.1% (n=142), %TIR 60.2%, 16.8%, 20-100%, daily insulin dose 0.89, 0.36, 0.16-2.05 (n=71).

Glycemic profiles were split according to whether any time was spent below glucose thresholds versus no time. Two glucose thresholds were considered: 3.0 mmol/L, corresponding to clinically significant hypoglycemia, respectively 3.9 mmol/L, corresponding to mild hypoglycemia. The number of glycemic profiles with any time spent below 3.0 mmol/L was smaller than the number with any time spent below 3.9 mmol/L (Table 1). MDG was significantly lower (Wilcoxon test p<0.0001) in those with both clinically significant and mild hypoglycemia compared to those without, while %CV was significantly higher (p<0.0001). There was no evidence of difference between %TIR between those with and without clinically significant hypoglycemia, however, %TIR was significantly higher in those with mild hypoglycemia compared to those without (Wilcoxon test p=0.01).

**Table 1.**
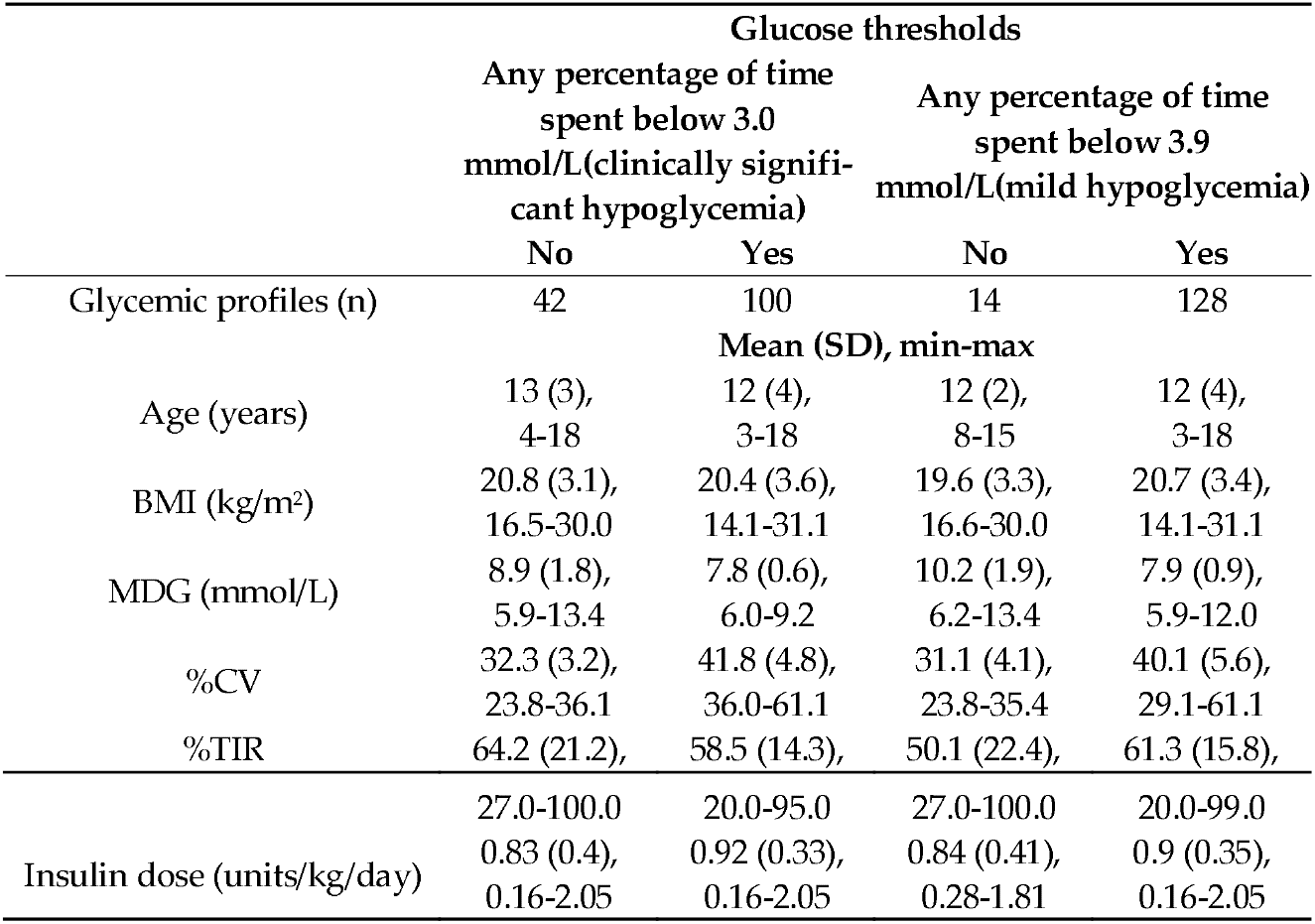
Glycemic profiles split by time spent below two glucose thresholds (3.0 and 3.9 mmol/L), together with mean and standard deviation (SD) age, BMI, MDG, %CV, TIR and insulin dose.

There was no evidence of difference between mean age, BMI or insulin dose between those with and those without hypoglycemia. All glycemic profiles with no percentage of time spent below 3.0 mmol/L had a %CV<36.1%, while all those with any time spent below had a %CV>36.0% (Table 1), indicating that, at least in our dataset, a threshold of 36.0% for %CV would almost perfectly split those with clinically significant hypoglycemia, from those without. We note, however, that this split was not as clean for mild hypoglycemia as the %CV for those with and without overlap in the interval (29.1%-35.4%). Similarly, there was an overlap when considering MDG for both clinically significant and mild hypoglycemia.

Considering one explanatory variable at a time, linear mixed models found statistically significant associations between explanatory variables MDG and %CV and response variables percentage of time spent below glucose thresholds 3.0 and 3.9 mmol/L (p-value Type II Wald chi-square test <0.0001 for all but MDG for clinically significant hypoglycemia with p=0.0005), and no evidence of association with age, BMI or insulin dose (p>0.2). Considering %TIR, there was no evidence of association with percentage of time spent below 3.9 mmol/L (p=0.8), however, there was a statistically significant association with percentage of time spent below 3.0 mmol/L (p=0.02).

Recursive Partitioning and Regression Trees which included both MDG and %CV were fitted for percentage of time spent below both glucose thresholds (Figure 1). For clinically significant hypoglycemia we also built trees which considered splitting on %TIR, however, there were no changes to the trees.

**Figure 1.**
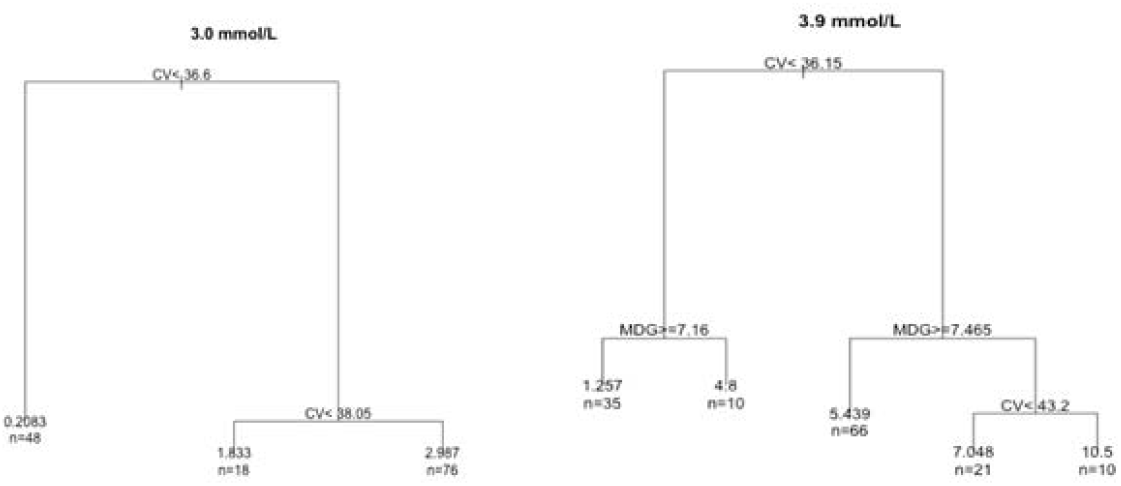
Regression trees with output features: percentage of time spent below the two glucose thresholds: 3.0 mmol/L (clinically significant hypoglycemia), respectively 3.9 mmol/L (mild hypoglycemia), and CV and MDG as the input features.

For both glucose thresholds a %CV of approximately 36% split the glycemic profiles with lowest percentage of time spent below the threshold. Interestingly, in clinical practice, for clinically significant hypoglycemia <1% of the time is often considered accepta-ble, while for mild hypoglycemia <4%, and these are also the splits identified by the trees (mean percentage of time, value of leaves 0.2% and 1.8%, respectively 4.8% and 5.4%). For clinically significant hypoglycemia, a further split on %CV separates the cases with an even higher percentage of time, noting however that even the most extreme percentages of time are fairly small in our dataset. For mild hypoglycemia, the second split is based on MDG, while those with the highest percentage of time spent below the 3.9 mmol/L threshold are separated by %CV values greater than 43.2%.

When considering classification trees for any percentage of time versus no time spent below the glucose thresholds, trees of depth 1 are chosen for both thresholds, with a split on %CV (36.1%) for clinically significant hypoglycemia, and on MDG (9.3 mmol/L) for mild hypoglycemia (Figure 2). That is, all patients with a %CV above 36.1% would be predicted to have had clinically significant hypoglycemia, while those with a %CV below 36.% would be predicted to have spent no time below 3.0 mmol/L. All patients with an MDG below 9.3 mmol/L would be predicted to have had mild hypoglycemia, while those with an MDG above 9.3 mmol/L would be predicted to have spent no time below 3.9 mmol/L.

**Figure 2.**
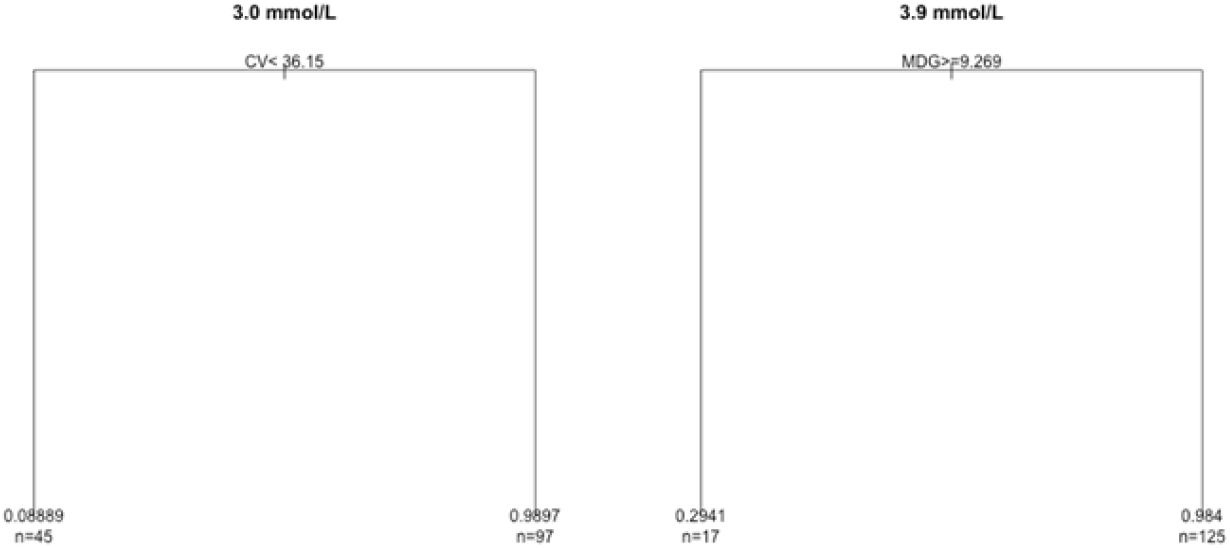
Classification trees with output features: any percentage of time versus no time spent below the two glucose thresholds: 3.0 mmol/L (clinically significant hypoglycemia), respectively 3.9 mmol/L (mild hypoglycemia), and CV and MDG as the input features.

Considering the relationship between %CV, respectively MDG, and the percentage of time spent below glucose thresholds, the percentage of time increased as %CV increased for both thresholds, and it decreased as MDG increased (Figure 3), regardless of the regression model considered.

**Figure 3.**
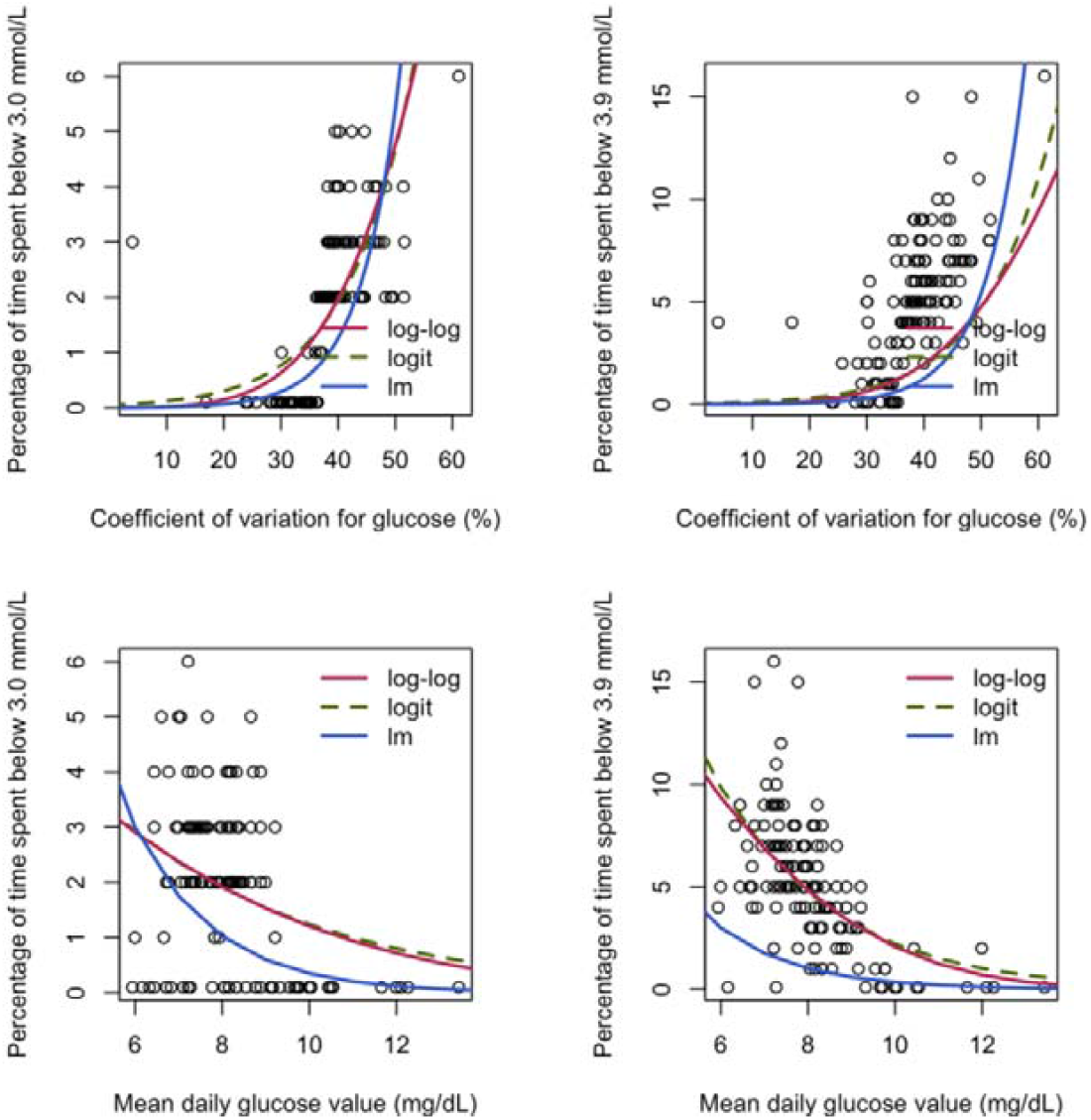
Predictions for the percentage of time spent below the two glucose thresholds 3.0 mmol/L, respectively 3.9 mmol/L from regression models using the coefficient of variation for glucose (%), respectively the mean daily glucose value (mg/dL) as explanatory variables. Models presented: Beta regression models using log-log and logit links, as well as from linear regression (lm) on the log-transformed data. Note: For all models percentages were divided by 100 to bring them between [0,1], and 0.001 was added to all response variables as Beta distribution does not include 0 and 1, and the natural logarithm of 0 is not defined. 0.001 was subtracted from the predictions

Considering the relationship between age category (<7, 7-13, >13 years) and any time spent below glucose thresholds: 3.0 mmol/L and 3.9 mmol/L, there was weak evidence (p=0.1, p=0.07) that there is a relationship between the two categorical variables (Table 2). The percentage of both clinically significant and mild hypoglycemia was highest in the <7 years age category, with similar percentages for 7-13 years and >13 years for clinical y significant hypoglycemia, but a lower percentage of mild hypoglycemia in 7-13 years compared to >13 years (Table 2).

**Table 2.**
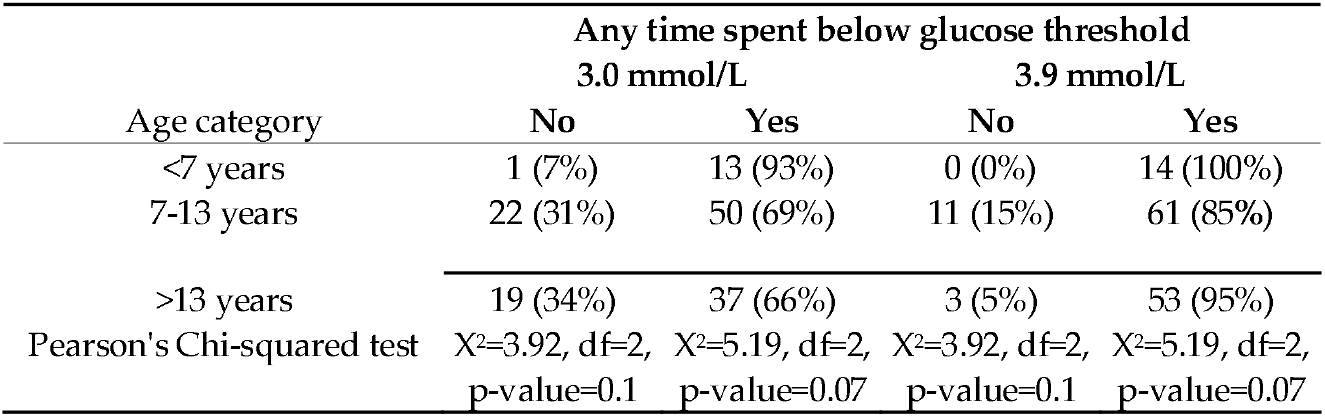
Contingency table for age category (<7, 7-13, >13 years) and any time spent below glucose thresholds: 3.0 mmol/L and 3.9 mmol/L, together with Pearson’s Chi-squared test to assess the relationship between these two categorical variables.

| Age category | Any time spent below glucose threshold |  |  |  |
| --- | --- | --- | --- | --- |
|  | 3.0 mmol/L |  | 3.9 mmol/L |  |
|  | No | Yes | No | Yes |
| <7 years | 1 (7%) | 13 (93%) | 0 (0%) | 14 (100%) |
| 7-13 years | 22 (31%) | 50 (69%) | 11 (15%) | 61 (85%) |

| >13 years | 19 (34%) | 37 (66%) | 3 (5%) | 53 (95%) |
| --- | --- | --- | --- | --- |
| Pearson's Chi-squared test | $\chi^2=3.92$, df=2,<br>p-value=0.1 | $\chi^2=5.19$, df=2,<br>p-value=0.07 | $\chi^2=3.92$, df=2,<br>p-value=0.1 | $\chi^2=5.19$, df=2,<br>p-value=0.07 |

## 4. Discussion

The results obtained in the present study highlight the key parameters predictive of hypoglycemic episodes among children and adolescents with T1DM. MDG was significantly lower (Wilcoxon test p<0.0001) in those with both clinically significant and mild hypoglycaemia compared to those without, while %CV was significantly higher (p<0.0001). The percentage of time spent below glucose thresholds increased as %CV increased for both thresholds, and it decreased as MDG increased. Regression trees found that for both glucose thresholds a coefficient of variation of approximately 36% split the glycemic profiles with lowest percentage of time spent below the threshold.

%CV and MDG were also found to be significantly associated with the presence of hypoglycemia in adults by Monnier et al., whose regression tree models identified %CV as the dominant variable for the 3.0 mmol/L threshold [34]. For the 3.9 mmol/L threshold MDG was identified as the dominant variable, however, their percentage of time spent below the thresholds was a lot more extreme than in our study, with our highest mean percentage of time spent below 3.9 mmol/L being 10.5 versus 37% in their regression tree. We also note that the values of their leaves at the MDG split were 11.4, respectively 14.1. The risk of clinically significant hypoglycemia was minimal when %CV values were below 36%, but increased exponentially as %CV rose and MDG decreased. One of the strengths of our analysis were the beta regression models, which are specifically designed to model continuous data bounded between 0 and 1, however, we also included an exponential fit for comparison with this study.

It was observed that in profiles with a %CV below 36.15% and MDG above 7.16 mmol/L, the mean time spent below the 3.9 mmol/L threshold was 4.8%, a value close to the 4% value recommended in American Diabetes Association guidelines [35]. This conclusion suggests that pediatric patients with lower glucose variability may achieve better glycemic control with a lower risk of hypoglycemia, consistent with findings reported by Urakami et al. [36].

According to our regression tree model results, an MDG value of 9.3 mmol/L was predictive of eliminating any time spent below the 3.9 mmol/L threshold. This MDG value exceeds the upper limit of euglycemia—7.8 mmol/L [37,38]—reinforcing the results from the specialized literature that show that the fear of hypoglycemia, often manifested through caregiver-driven overcorrection of glucose levels, may contribute to suboptimal glycemic control, particularly in the pediatric population [39–41].

Until recently, only a small proportion of individuals with T1DM were able to simultaneously achieve such targets—low %CV and near-normal MDG concentration—using conventional insulin therapy, whether through multiple daily injections or CSII using an insulin pump. Several factors have been implicated, one of which is pediatric age itself, which appears to play an important role in modulating the frequency and severity of hypoglycemic episodes.

Clinically significant hypoglycemia has been associated with age under 5 years, with the likelihood of developing hypoglycemia being approximately six times higher in this age group compared to children over 10 years [42]. In the studied cohort, patients younger than 7 years demonstrated the highest frequency of both mild and clinically significant hypoglycemic episodes. This observation is of particular importance, given that recurrent and early exposure to hypoglycemia predisposes patients to maladaptive physiological responses.

Since the iatrogenic development of impaired hypoglycemia awareness and autonomic failure are considered mediators of severe hypoglycemia, medical efforts to manage both mild and clinically significant hypoglycemic episodes should be prioritized—especially in pediatric patients, whose age at disease onset has shown concerning trends in recent years [43–45]. Furthermore, evidence from large-scale clinical trials, real-world studies, and meta-research supports the association between hypoglycemia and adverse cardiovascular outcomes in patients with diabetes, reinforcing the long-standing principle attributed to Erasmus of Rotterdam, “prevention is better than cure,” and underscoring the importance of preventing this complication from early childhood [46–49].

Integrating stable clinical factors (such as age at evaluation) with dynamic indicators of glycemic control (%CV; MDG) enables predictive and proactive stratification of pediatric patients at increased risk for recurrent and clinically significant hypoglycemia. Future research should explore how glycemic variability and mean daily glucose contribute to fear of hypoglycemia experienced by both patients and caregivers. Accordingly, the development and rigorous evaluation of psychotherapeutic interventions tailored to the pediatric population are essential.

Several limitations must be acknowledged. The study has an observational design and a relatively short monitoring period for AGP profiles. Additionally, rtCGM sensors may occasionally have been inaccurate at lower glucose ranges. The study cohort consisted of pediatric patients using rtCGM devices whose hypoglycemia alarms could not be disabled due to young age, limited communication abilities, and caregiver fear of hypoglycemia.

In conclusion, the findings of our study provide a clear answer to the proposed research question by demonstrating that glycemic variability (assessed through the coefficient of variation), MDG levels, and patient age at evaluation are key factors in predicting and preventing hypoglycemia risk. Based on these results, achieving and maintaining a %CV below 36% may serve as a primary target within strategies aimed at reducing the incidence of hypoglycemic episodes in children and adolescents with type 1 diabetes.

## Supplementary Materials

None.

## Author Contributions

Conceptualization, A.M.S and C.D.D; methodology, A.M.S; formal analysis, A.M.S, L.D; investigation, A.M.S and A.R; resources, A.M.S and C.D.D; data curation, A.M.S.; writing—original draft preparation, A.M.S and C.D.D.; writing—review and editing, AM.S.; visualization, L.D and A.R.; supervision, C.D.D.; project administration, A.M.S, C.D.D and L.D. All authors have read and agreed to the published version of the manuscript.

## Institutional Review Board Statement

This study was conducted according to the guidelines of the Declaration of Helsinki and approved by the Institutional Review Board (or Ethics Committee) of the Emergency County Hospital of Oradea (protocol number 19384/ 26 June 2025) and University “Lucian Blaga” Sibiu (No.57/12 November 2025).

## Informed Consent Statement

Individual patient consent was not required due to the retrospective nature of this study. However, the legal representatives of all participating patients provided gen-eral written consent for the processing and utilization of medical data for scientific and research purposes upon hospital admission.

## Data Availability Statement

The datasets presented in this article are not readily available because they belong to the Department of Diabetology and Nutrition Diseases of Oradea County Clinical Emergency Hospital. Requests to access the datasets should be directed to the corresponding author.

## Acknowledgments

This work uses data provided by patients and collected by the Department of Diabetology and Nutrition Diseases of Oradea County Clinical Emergency Hospital as part of their care and support. We thank all the people who contributed to the database.

## Abbreviations

The following abbreviations are used in this manuscript:

CGM: continuous glucose monitoring
CV: coefficient of variation
MDG: mean daily glucose
T1DM: Type 1 diabetes mellitus
CSII: continuous subcutaneous insulin infusion
AGP: Ambulatory Glucose Profile
rtCGM: real-time
CGM: TIR time in range
BMI: body mass index
IQR: interquartile range
SD: standard deviation

## Disclaimer/Publisher’s Note

The statements, opinions and data contained in all publications are solely those of the individual author(s) and contributor(s) and not of MDPI and/or the editor(s). MDPI and/or the editor(s) disclaim responsibility for any injury to people or property resulting from any ideas, methods, instructions or products referred to in the content.

